# Monitoring sexual hormones in women going to high altitude – a pilot study

**DOI:** 10.1101/2024.12.04.24318483

**Authors:** Aijan Taalaibekova, Michelle Meyer, Stefanie Ulrich, Gulzada Mirzalieva, Maamed Mademilov, Mona Lichtblau, Cornelia Betschart, Talant M. Sooronbaev, Silvia Ulrich, Konrad E. Bloch, Michael Furian

## Abstract

**Background:** The susceptibility to acute mountain sickness (AMS) in relation to sexual hormones in women remains elusive, partly because hormones could not be conveniently measured. We evaluated a novel kit for self-monitoring sexual hormones in women and recorded AMS incidence during high-altitude sojourns.

**Methods:** Two groups of healthy, premenopausal women, mean±SD age 23.1±2.3 y, residing <1,000m underwent baseline evaluations at 760 m before travelling to and staying for 2 days and nights (48 hours) at 3,100 m or 3,600 m, respectively. Participants self-monitored morning urine sexual hormone concentrations (estrone-1-glucuronide, E1G, pregnanediol-3-alpha-glucuronide, PdG, and luteinizing hormone, LH) daily for 30d including altitude sojourns using the simple *“Proov”* kit (MFB Fertility Inc). Follicular and luteal menstrual cycle phases detected by LH peak, altitude-related adverse health effects (ARAHE), AMS (Lake Louise score 2018 [LLS] ≥ 3 points including headache) and pulse oximetry (SpO_2_) were assessed.

**Results:** 1,172 of 1,250 (93.8%) hormone measurements were successful, 78 of 1,250 (6.2%) failed due to nonadherence or technical failure. At 3,600 m, mean differences in urinary PdG concentration were 3.8 mcg/ml (95%CI, 0.6 to 7.1) between luteal and follicular cycle phases. At 3,100 m, corresponding difference was 8.5 mcg/ml (95%CI, 5.0 to 12.0). At 3,100 m, 9 of 21 (43%) women were diagnosed with AMS with SpO_2_ of 93.0±1.6% and LLS of 0.3±1.4 in the morning after the first night. At 3,600 m, 12 of 21 (57%) women had AMS (p=0.355 versus 3,100 m) with SpO_2_ of 86.8±1.8% (p<0.05 vs. 3,100 m) and LLS of 1.9±1.4 (p<0.05 vs. 3,100 m).

**Conclusion:** Self-monitoring female sexual hormones during high-altitude field studies with the employed kit is feasible and provides physiologically plausible trends of hormone levels over the menstrual cycle. Our data provide a valuable basis for designing further studies to evaluate AMS susceptibility in women.

## Introduction

To date, there is no reliable, validated, and simple tool for monitoring hormonal concentrations or reliably identifying menstrual cycle phases in women during altitude sojourns. The validation of a reliable tool is particularly important since the pathophysiology of altitude-related adverse health effects (ARAHE), particularly acute mountain sickness (AMS) and factors for individual susceptibility remain elusive (Imray et al., 2010). Importantly, it has not yet been quantitatively assessed whether women are more, equally or less prone than men to develop AMS men (Derstine et al., 2023a). The unresolved issue might originate from underrepresentation of women in high altitude studies, lack of powered sex-specific analyses or the poor characterization of premenopausal women in regard of the menstrual cycle phase, sex hormone concentrations and cycle day while staying at high altitude. Hormonal changes related to the menstrual cycle phases can alter breathing control, which may result in different altitude-related adaptations. Progesterone, which is up-regulated during the luteal phase, is a well-known respiratory stimulant associated with elevated minute ventilation and decreased PaCO_2_ during the luteal phase compared to the follicular phase (Behan and Wenninger, 2008; Richalet et al., 2020). Despite these observations at low altitude, previous studies found no relationship between menstrual cycle phase or sex hormones and AMS susceptibility, however, both studies did not quantify and assess sex hormones and cycle phase at high altitude. In contrast, a small study conducted in 1997 and 1998 assessed sex hormone levels at high altitude and found a significant difference in AMS incidence of 15% between menstrual cycle phases, though it concluded that the difference was not clinically relevant. Based on these previous reports and technological advancements, the primary purpose of this prospective pilot study was to evaluate the tolerability of using a commercially available fertility self-monitoring kit to measure urinary sex hormone concentrations and to determine the menstrual cycle phases and cycle days when travelling to high altitude. Additionally, to enable sample size estimations and provide preliminary insights into potential differences in clinical and physiological outcomes between the luteal and follicular menstrual cycle phases, additional outcomes such as AMS, nocturnal pulse oximetry, subjective sleep quality and vital signs were assessed.

## Methods

### Study design and setting

This prospective, non-randomized pilot study was conducted at the National Center of Cardiology and Internal Medicine, Respiratory Medicine Department, Bishkek (760 m), Kyrgyzstan; at the Swiss-Kyrgyz High-Altitude Clinic in Tuja Ashu (3,100 m), Kyrgyzstan, which is a 4 hour bus ride away from the first location and where participants spent 2 days and nights (48 hours); and at the High Altitude Kumtor Gold Mine operation facility (3,600 m), Kumtor, Kyrgyzstan, 8 hours by bus, where participants also spent 2 days and nights (48 hours). The ascent to 3,100 m (Expedition 1, n=21) was performed in July 2022; the ascent to 3,600 m (Expedition 2, n=21) in September 2022 and both altitude ascents were scheduled independently of the menstrual cycle phases or cycle days of the participants. Sixteen participants from Expedition 1 participated also in Expedition 2 and 5 new participants had to be recruited (**Figure 1**).

**Figure 1.**
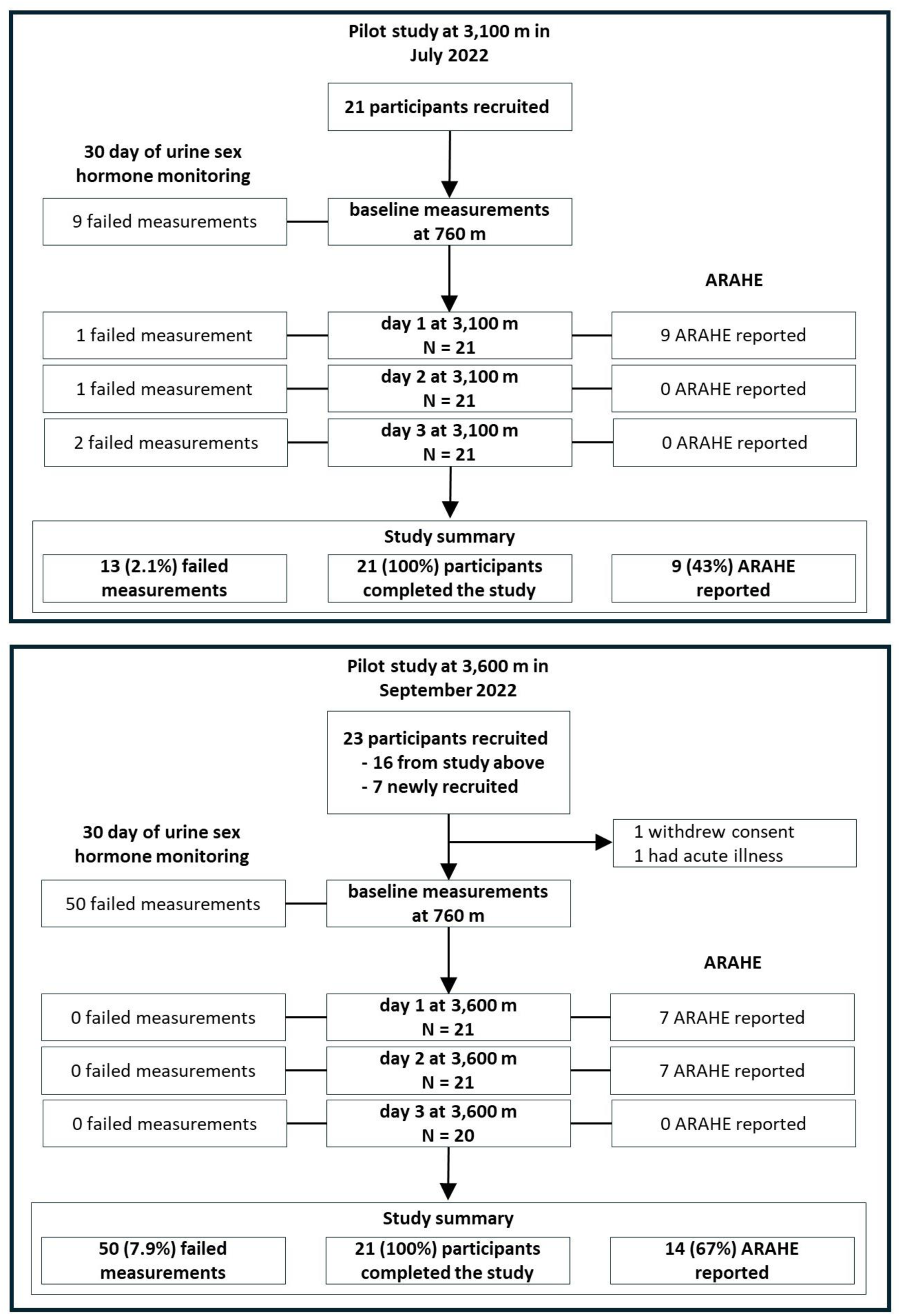
Study design of the pilot study. A pilot study validating the commercially available fertility monitor *Proov* to monitor hormones and menstrual cycle phases before, during and after an altitude sojourn. Women monitored their hormone for 30 consecutive days. During the hormone monitoring they ascended and stayed for 2-day and nights (48 hours) at either 3,100 (Expedition 1) or 3,600 m (Expedition 2), respectively.

Participants in this study were recruited through advertisements at different universities around Bishkek. Included were premenopausal, eumenorrheic, non-smoking, healthy women with a BMI>18 kg/m^2^ and <30 kg/m^2^, aged 18 to 44 years, living at altitudes under 1,000 m. Exclusion criteria were any pre-existing disease, regular intake of medication (including oral contraception), other types of hormonal contraceptives (hormonal intrauterine device, vaginal ring, subcutaneous injections or implants), pregnancy or nursing, and any altitude trip <4 weeks prior to the study. The study was approved by the Ethic Committee of the National Center of Cardiology and Internal Medicine named after academician M. Mirrakhimov (Protocol No. 07 in the year of 2022) and participants provided written informed consent.

### Hormone monitoring

All participants monitored urine metabolite concentrations of estrogen, luteinizing hormone, and progesterone for 30 consecutive days using two different sex hormone monitoring devices, described in detail below.

#### Proov Full Cycle Hormone Insights Kit (Proov, MFB Fertility Inc., CO, US)

This device was used during both Expedition 1 & 2. The procedure requires a hormone test strip into a fresh morning urine sample for 10 –20 seconds, collected before any food or fluid intake. The *Proov* multi-hormone test strip lateral flow assay uses gold nanoparticles and buffered sample pads to adjust for pH and hydration levels, filter unwanted particulates and bind contaminants in urine that may interfere with test accuracy. The strip contains three test lines and one control line, corresponding to estrone-1-glucuronide (E1G), pregnanediol-3-alpha-glucuronide (PdG), and luteinizing hormone (LH) (beta subunit). After 10 minutes, the participant takes a photo of the urine test strip with the smartphone and uploads it onto the company server *using the Proov* app. The application server employs machine learning algorithms specifically tailored to analyze photographed images of the test strip, accounting for variations in camera, lighting, and operating system, while also verifying input and output regularities and mathematically deriving the associated hormone levels (Wegrzynowicz et al., 2022). In the case of internet connection loss, the company can conduct offline analysis if the time, day and picture of the test strip of the participant are provided. In this study, daily concentrations of sex hormones E1G, PdG and LH were extracted from the *Proov* app.

#### *Mira* Fertility Tracking Kit (*Mira* – Crow Canyon PL, San Ramon, CA, USA)

This device was used during Expedition 2. Identical to the *Proov* testing procedure, Mira analysis requires to dip a hormone test strip for 10 –20 seconds into a fresh urine sample collected in the morning before any fluid or food intake. Thereafter, the strip will be inserted into the *Mira* device to initiate the quantitative analysis of estrogen-3-glucuronide (E3G), LH and PdG. After approximately 21 minutes the results will be synchronized with the *Mira* smartphone app through Bluetooth. In case of an analysis error, the strip cannot be re-analyzed.

Together with the information regarding the first day of the participant’s last menstrual bleeding, the measured sex hormones were visually plotted. Based on the visually confirmed LH peak and increment of PdG, the follicular (first day of menstrual bleeding to LH peak) and luteal menstrual cycle phases (LH peak to end of cycle) were identified.

### Medical history, symptoms, and clinical examination

A medical history has been obtained, including information on menstrual cycle length, and the starting and ending days of the participants’ last menstruation. Physical examination included measurements of weight, height, blood pressure, heart rate, and cardiac and pulmonary auscultation. At altitudes, adverse events and AMS incidences were assessed, using both the revised and original Lake Louise questionnaires (LLQ), as well as the Environmental Symptom Questionnaire cerebral score (AMSc) (Roach et al., 2018; Sampson et al., 1983). Primary definition of AMS incidence was a 2018 LLQ score of ≥3 points including headache, over the two-day period at high altitude; secondary AMS definitions included a 1993 LLQ score of ≥3 points and an AMSc of ≥0.7. Since previous studies in both sick and healthy persons traveling to high altitudes reported the occurrence of non-AMS related ARAHE (Furian et al., 2022), this pilot study additionally assessed the incidence of ARAHE at high altitudes. ARAHE was defined as any medical health condition (including AMS) that occurred at high altitude and required medical intervention or premature descent.

Nocturnal pulse oximetry was performed at 760 m and at 3,600 m to assess nocturnal oxygenation. Sleepiness was assessed with the Karolinska Sleepiness Scale and subjective sleep quality with a 100-mm visual analogue scale ranging from 0 (extremely poor) to 100 (excellent) (Kaida et al., 2006).

### Outcomes

The main outcomes included the proportion and comparison of successful applications of the *Proov* and *Mira* hormone test kits, their failure rates, and reasons for failure. Additional outcomes included daily hormone concentrations throughout the menstrual cycle, differences in estrogen and progesterone concentrations between the follicular and luteal menstrual cycle phases, and the timing of the LH peak, defined as the day with the highest median value of 3 consecutive daily concentrations. The incidence of AMS and other ARAHE, nocturnal pulse oximetry, and subjective sleep quality as well as physiological variables in the different menstrual cycle phases were determined.

### Statistics

No sample size estimation was performed for this pilot study. A convenience sample of 20 participants per expedition to 3,100 m and 3,600 m were recruited. The feasibility of hormone monitoring at low and high altitudes, as well as during experimental field expeditions, was quantified using descriptive statistics on successful instances of hormone monitoring, hormone concentrations, and failure rates associated with the use of the *Proov* or *Mira devices*, respectively. Based on the findings of the two devices, menstrual cycle phases, ovulation day, and failure rates were compared using Chi-square test statistics and Pearson correlation. AMS incidences and physiological variables were then compared between menstrual cycle phases and between altitudes using mixed linear regressions to compute mean differences and 95% confidence intervals. Continuous outcomes are presented as means ± SE and binary outcomes as numbers and proportions. Two exploratory regression models were used to evaluate 1) the associations between sex hormone concentrations and SpO_2_ (mixed linear regression model); 2) the associations between menstrual cycle phases and the incidence of AMS (logistic regression model). Due to the small sample size, the parameters for the mixed models were predefined. Therefore, in the model for SpO_2_, sex hormones (E1G, LH and PdG) and assessment altitudes (760 m, 3,100 m and 3,600 m) were used as predictors. A two-sided p-value of 0.05 was considered statistically significant. Statistical analyses were performed in STATA version 15 (StataCorp LLC, College Station, Texas, USA).

### Results

The study flow chart is presented in **Figure 1**. Out of 26 participants with a mean ± SD age of 23.5±2.5 years, a total of 21 (80.7%) went to 3,100 m; 21 (80.7%) went to 3,600 m and 16 (61.5%) participants completed both expeditions (**Table 1**). The menstrual cycle length assessed with the *Proov* app was 29±2 days and almost identical with what participants reported during the screening visit (30±2 days). The maximum deviation between the assessed and reported menstrual cycle length was 9 days.

**Table 1:** Participant demographics

| Variable | All | Participants going to 3,100 m | Participants going to 3,600 m | Participants going to 3,100 and 3,600 m |
| --- | --- | --- | --- | --- |
| N | 26 | 21 | 21 | 16 |
| Age, years | 23.5 ± 2.5 | 23.1 ± 2.3 | 23.7 ± 2.7 | 23.1 ± 2.5 |
| Height, m | 1.63 ± 0.07 | 1.61 ± 0.07 | 1.63 ± 0.08 | 1.61 ± 0.07 |
| Weight, kg | 59.3 ± 8.2 | 59.0 ± 7.6 | 59.9 ± 7.5 | 60.3 ± 8.0 |
| BMI, kg/m<sup>2</sup> | 22.4 ± 3.2 | 22.7 ± 3.3 | 22.7 ± 3.0 | 23.3 ± 3.5 |
| **Menstrual Cycle** |  |  |  |  |
| Historical Cycle length, days<sup>1</sup> | 30 ± 3 | 29 ± 1 | 30 ± 5 | 29 ± 1 |
| Measured Cycle length, days<sup>2</sup> | 29 ± 2 | 29 ± 2 | 29 ± 2 | 29 ± 2 |
| Length of menstruation, days<sup>1</sup> | 5 ± 2 | 6 ± 1 | 5 ± 2 | 6 ± 1 |
| Proov hormone monitoring measurement errors, N | 78 of 1,250 (6.2%) | 28 of 620 (4.5%) | 50 of 630 (7.9%) | 59 of 952 (6.2%) |
| Mira hormone monitoring measurement errors, N | 71 of 624 (11.4%) | NA | 71 of 624 (11.4%) | NA |

Values presented in mean ± SD. BMI, body mass index. <sup>1</sup>Provided by asking each participant during the screening visit. <sup>2</sup>Assessed using the *proov* Fertility app.

#### Hormone monitoring and comparisons between menstrual cycle phases

When using the *Proov* hormone test kit, the daily urinary hormone measurement error rates were 4.5% (95%CI of 3.0 to 6.5%) in the group travelling to 3,100 m, 7.9% (95%CI of 5.9 to 10.3%) in those travelling to 3,600 m, and 6.2% overall (95%CI of 5.0 to 7.7%) (**Table 1**). At 3,600 m, the corresponding error rate when using the *Mira* hormone test kit was 12.2% (95%CI of 9.8 to 15.0%, P<0.001 between error rates of *Proov* vs. *Mira* at 3,600 m). The urinary hormone monitoring and the assignment of the participants to a specific menstrual cycle phases and cycle day while staying at altitude were successful and are illustrated in **Figure 2, Upper Panel**. **Figure 2, Lower Panel** shows the proportions of participants suffering from AMS during the follicular compared to the luteal phase, however, exploratory mixed logistic regression analysis showed no association between menstrual cycle phases and the incidence of AMS defined by the 2018 LLQ criteria (Odds ratio of 0.50, 95%CI 0.09 to 2.8, p=0.430). **Table 2** shows the comparison of the menstrual cycle phases and the mean differences (95%CI) in other physiological and clinical outcomes between the luteal and follicular phases. Both progesterone and estrogen were higher during the luteal compared to the follicular phase. During the nights at 3,600 m, participants in the luteal phase showed a trend toward faster SpO_2_ acclimatization between nights 1 and 2, with a mean difference (95%CI) of 2.1% (−0.3 to 4.6), p = 0.085, compared to participants in the follicular phase (**Table 3**). These findings align with the results from mixed linear regression analysis on daytime SpO_2_, showing a positive correlation with PdG concentration and a negative correlation with E1G and absolute altitude of measurement (**Supplement Table S2**).

**Figure 2.**
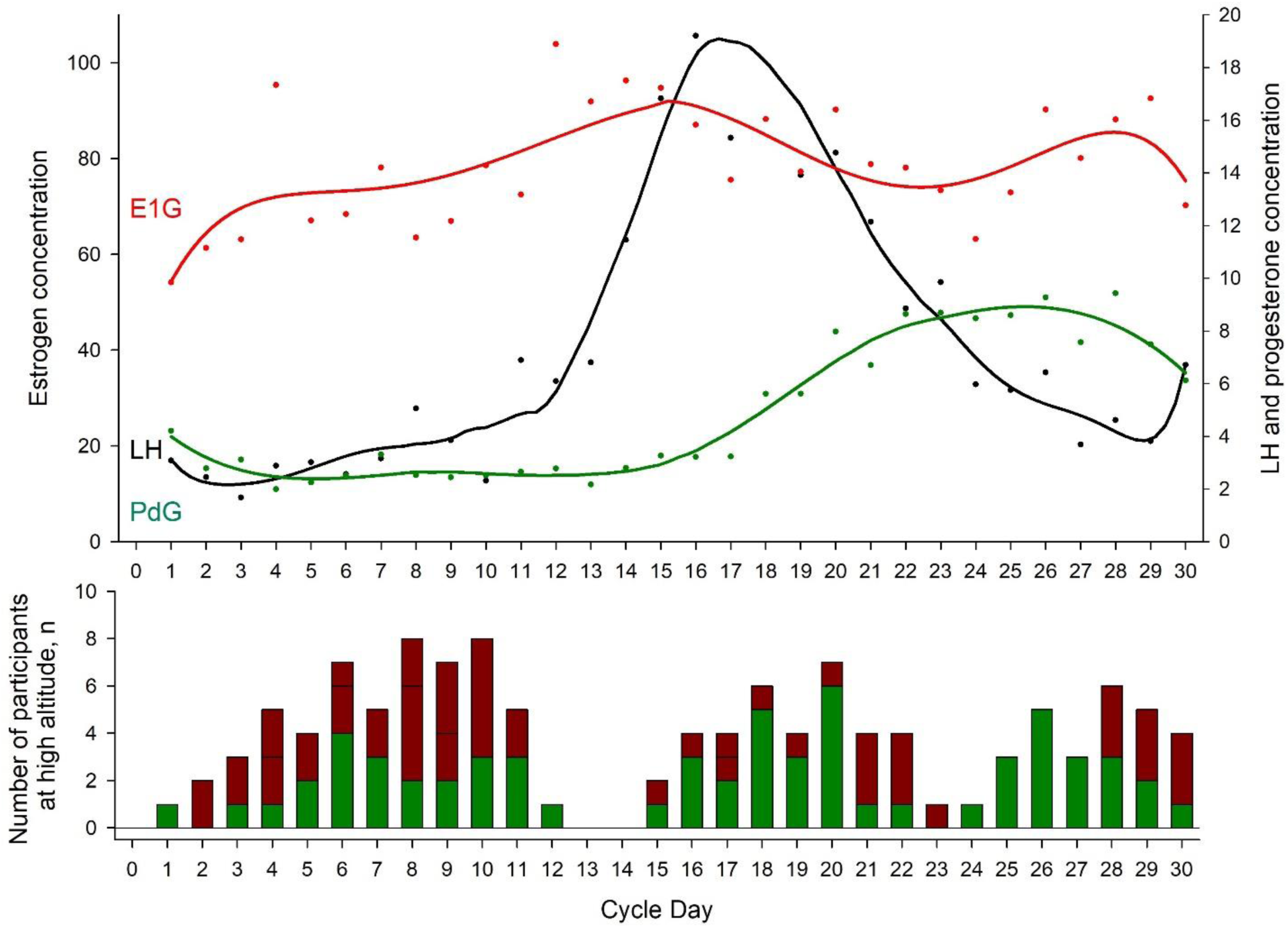
Hormone profile and acute mountain sickness incidence. **Upper panel** shows the mean hormone concentrations assessed by *Proov* for each menstrual cycle in coloured dots. The connecting line represents a smoother of the corresponding hormone. This figure indicates a commonly seen hormone profile of oestrogen (E1G), progesterone (PdG) and luteinizing hormone (LH) and confirms the feasibility of assessing hormones by urine stripes. The follicular phase was defined as the time from the first day of menstrual bleeding to the LH peak (Cycle Day 16 in the figure), while the luteal phase was defined as the time from the LH peak to the day before the next menstrual bleeding. **Lower panel** shows the absolute number of participants staying at altitude, stratified by menstrual cycle days. The green coloured proportion of the bars represent the number of participants without acute mountain sickness (acute mountain sickness was defined as a 2018 Lake Louise questionnaire score of ≥3 points including headache.); the red proportion of the bars represent the number of participants with, or the days after the diagnosis of acute mountain sickness. For example, when a participant experienced acute mountain sickness on day 2 at 3,600 m, this person was medically treated and, if feasible, descended to low altitude on day 3, however, day 2 and 3 were marked as AMS-positive in this figure.

**Table 2:** Insights into menstrual cycle phase-related differences at high altitude.

| Variable | Group 1 at 3,100 m |  |  | Group 2 at 3,600 m |  |  |
| --- | --- | --- | --- | --- | --- | --- |
|  | Follicular phase | Luteal phase | Mean MCP difference (95% CI) | Follicular phase | Luteal phase | Mean MCP difference (95% CI) |
| N | 12 | 7 |  | 11 | 10 |  |
| **Hormones** |  |  |  |  |  |  |
| Estrogen, ng/ml | 85 ± 20 | 92 ± 26 | 6 (-57 to 70) | 81 ± 22 | 145 ± 22 | 64 (5 to 123) |
| Progesterone, µg/ml | 1.6 ± 1.1 | 10.1 ± 1.4 | 8.5 (5.0 to 12.0) | 5.2 ± 1.2 | 9.0 ± 1.2 | 3.8 (0.6 to 7.1) |
| Luteinizing hormone, mIU/ml | 4.6 ± 3.1 | 3.4 ± 4.1 | -1.2 (-11.3 to 8.9) | 7.7 ± 3.4 | 10.2 ± 3.4 | 2.6 (-6.9 to 12.0) |
| **Clinical examination** |  |  |  |  |  |  |
| Systolic blood pressure, mmHg | 106 ± 3 | 108 ± 3 | 2 (-4 to 8) | 104 ± 2 | 106 ± 2 | 2 (-4 to 8) |
| Diastolic blood pressure, mmHg | 75 ± 2 | 71 ± 2 | -3 (-9 to 3) | 77 ± 2 | 75 ± 2 | (-2 (-8 to 4) |
| Heart rate, bpm | 88 ± 3 | 87 ± 4 | 0 (-10 to 9) | 96 ± 3 | 95 ± 3 | -1 (-9 to 8) |
| SpO<sub>2</sub>, % | 92.6 ± 0.5 | 93.9 ± 0.6 | 1.3 (-0.3 to 2.9) | 86.3 ± 0.6 | 86.6 ± 0.5 | 0.3 (-1.3 to 1.8) |
| **Acute mountain sickness** |  |  |  |  |  |  |
| 2018 Lake Louise Score | 0.4 ± 0.3 | 0.3 ± 0.5 | -0.1 (-1.2 to 1.0) | 2.2 ± 0.4 | 1.8 ± 0.4 | -0.4 (-1.4 to 0.6) |
| 1993 Lake Louise Score | 0.5 ± 0.5 | 0.3 ± 0.6 | -0.2 (-1.8 to 1.4) | 3.0 ± 0.5 | 2.5 ± 0.5 | -0.5 (-2.0 to 1.0) |
| AMSc score | 0.06 ± 0.08 | 0.03 ± 0.11 | -0.04 (-0.30 to 0.22) | 0.48 ± 0.09 | 0.30 ± 0.09 | -0.18 (-0.42 to 0.06) |
| **Subjective sleep quality** |  |  |  |  |  |  |
| Karolinska Sleepiness Scale | 4.1 ± 0.4 | 3.9 ± 0.5 | -0.2 (-1.4 to 1.1) | 4.4 ± 0.4 | 3.6 ± 0.4 | -0.8 (-2.0 to 0.4) |
| Time until falling asleep, min | 23 ± 12 | 21 ± 16 | -2 (-42 to 38) | 70 ± 14 | 45 ± 14 | -25 (-62 to 13) |
| Number of nocturnal awakenings, n | 1 ± 0 | 1 ± 0 | 0 (-2 to 1) | 3 ± 0 | 1 ± 0 | -2 (-3 to 0) |
| Waketime after sleep onset, min | 5 ± 9 | 2 ± 12 | -2 (-32 to 28) | 11 ± 10 | 37 ± 10 | 26 (-3 to 54) |
| Subjective sleep quality, % | 71 ± 5 | 79 ± 6 | 8 (-7 to 22) | 61 ± 5 | 51 ± 5 | -10 (-24 to 4) |

Values presented as means ± SE or mean differences (95%CI). MCP, menstrual cycle phase; SpO<sub>2</sub>, arterial oxygenation assessed by finger pulse oximetry; AMSc, Environmental Symptom Questionnaire cerebral score. No statistical analyses were performed due to the nature of a pilot study.

**Table 3:**
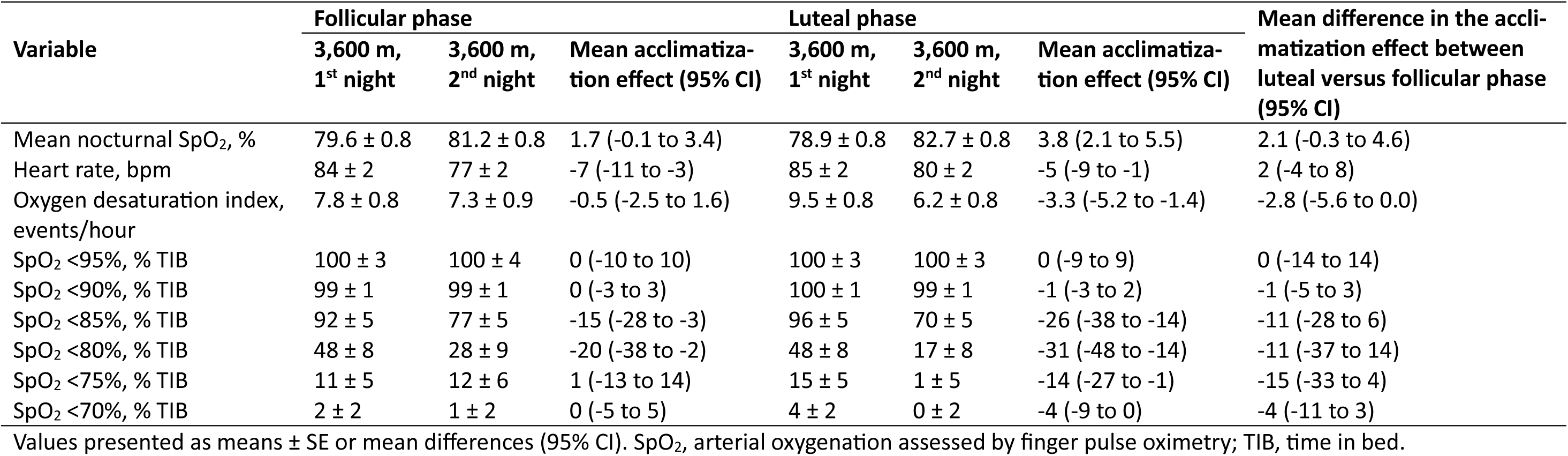
Nocturnal pulse oximetry stratified by menstrual cycle phases.

#### AMS and other ARAHE

During the altitude sojourns, 9 of 21 (43%) participants at 3,100 m and 12 of 21 (57%) at 3,600 m developed AMS (**Table 4**, p=0.355 between altitudes). At 3,600 m, 24% of participants experienced other ARAHEs not related to AMS, including fainting (N=1) and fever (N=1) on the day of arrival; panic attack (N=1) during the first night and abdominal pain (N=1) and diarrhea (N=1) over the time course of the 2 days and nights (**Table 4**). As shown in **Figure 3**, a high proportion of participants who experienced an ARAHE or AMS at 3,100 m also experienced one at 3,600 m.

**Table 4:** AMS and other ARAHE incidence rates.

|  | 3100 m (n=21) | 3600 m (n=21) | p-value |
| --- | --- | --- | --- |
| AMS, 2018 LLS $\geq 3$ | 9 (43%) | 12 (57%) | 0.355 |
| AMS, 1993 LLS $\geq 3$ | 9 (43%) | 12 (57%) | 0.355 |
| AMS, AMSc $\geq 0.7$ | 1 (5%) | 6 (29%) | 0.093 |
| **Composite endpoint** |  |  |  |
| ARAHE incidence, total* | 9 (43%) | 14 (67%) | 0.121 |
| **Components of ARAHE** |  |  |  |
| AMS, LLS 2018 $\geq 3$ or AMSc $\geq 0.7$ | 9 (43%) | 13 (62%) | 0.217 |
| Fainting | 0 (0%) | 1 (5%) | 1.000 |
| Fever | 0 (0%) | 1 (5%) | 1.000 |
| Panic attack | 0 (0%) | 1 (5%) | 1.000 |
| Abdominal pain | 0 (0%) | 1 (5%) | 1.000 |
| Diarrhea | 0 (0%) | 1 (5%) | 1.000 |
| Vomiting | 0 (0%) | 3 (14%) | 0.232 |

\*Participants experienced one or more ARAHE. ARAHE, altitude-related adverse health effect; AMS, acute mountain sickness; LLS, Lake Louise Questionnaire score; AMSc, Environmental Symptom Questionnaire cerebral score.

**Figure 3.**
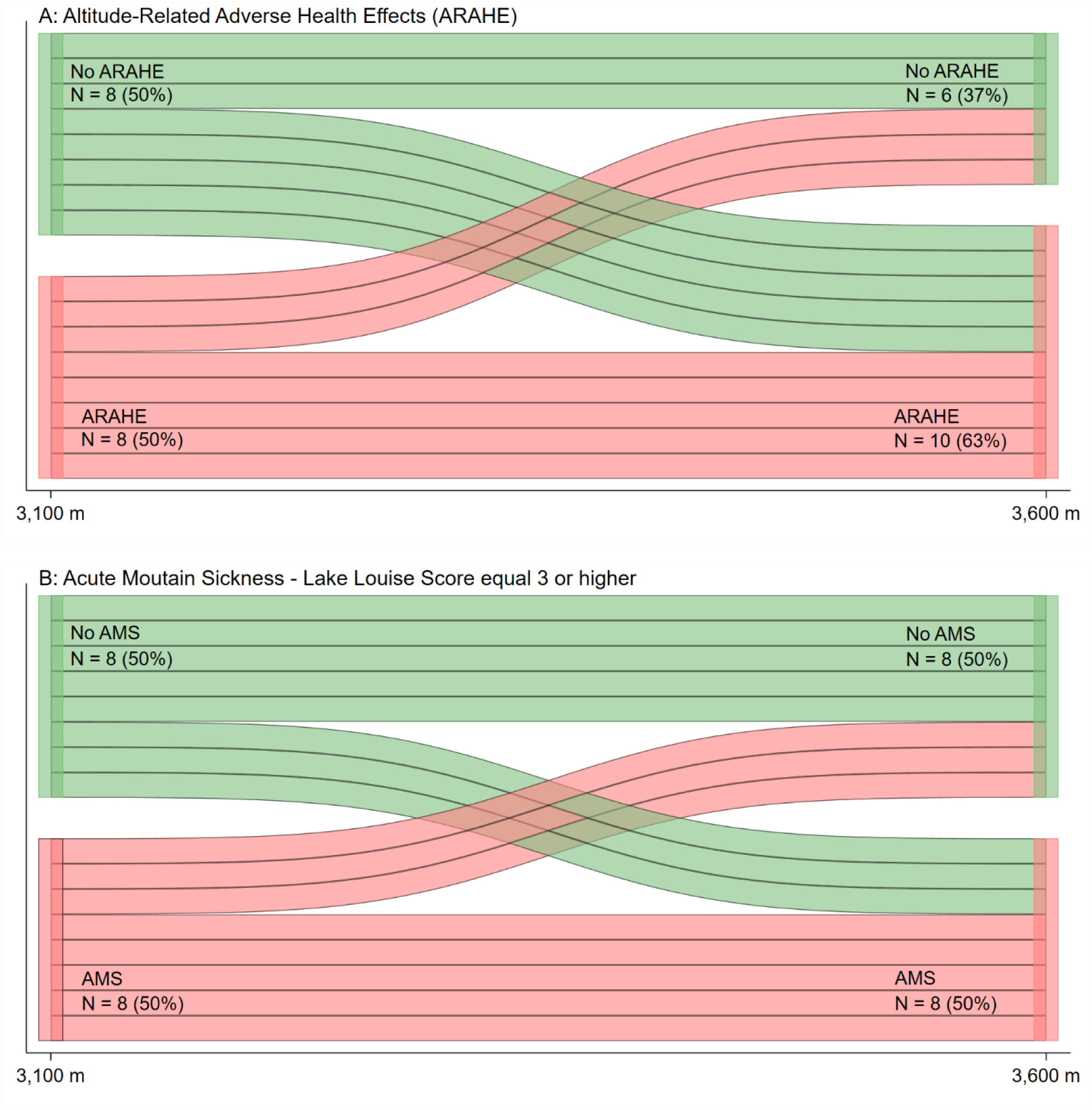
Incidences of altitude-related adverse health effects (ARAHE) and acute mountain sickness (AMS) in participants travelling to 3,100 and 3,600 m (N = 16). The data from 16 participants who completed both expeditions were analyzed. Green bars indicate participants free of ARAHE **(A)** or AMS **(B)** while red bars indicate participants suffering from ARAHE or AMS at the respective altitude. Acute mountain sickness was defined as a 2018 Lake Louise questionnaire score of ≥3 points including headache.

#### Secondary outcomes

On the morning of day 2 at 3,100 m, no significant differences were observed in vital parameters nor subjective sleep quality compared to 760 m, except for an expected decrease in SpO_2_ **(Table 5)**. In contrast, at 3,600 m, a significant decrease in SpO_2_ and subjective sleep quality as well as a significant increase in diastolic blood pressure, heart rate and AMS severity were observed (**Table 5**). At 3,600 m, nocturnal SpO_2_ dropped and oxygen desaturation index increased during the first night compared to 760 m, but significantly improved during the second night (**Supplement Table S1**).

**Table 5:** Secondary findings assessed in the morning on day 2

| Variable | Group 1 |  |  | Group 2 |  |  | Mean difference<br>in altitude effect<br>(95% CI) |
| --- | --- | --- | --- | --- | --- | --- | --- |
|  | 760 m | 3,100 m | Mean altitude<br>effect (95% CI) | 760 m | 3,600 m | Mean altitude<br>effect (95% CI) |  |
| **Clinical examination** |  |  |  |  |  |  |  |
| Systolic blood pressure, mmHg | 106 ± 2 | 106 ± 2 | 0 (-3 to 3) | 105 ± 2 | 105 ± 2 | 1 (-2 to 4) | 1 (-4 to 5) |
| Diastolic blood pressure, mmHg | 73 ± 2 | 74 ± 2 | 1 (-3 to 5) | 70 ± 1 | 76 ± 2 | 6 (2 to 9)* | 5 (0 to 10) |
| Heart rate, bpm | 85 ± 2 | 87 ± 2 | 2 (-3 to 7) | 78 ± 2 | 95 ± 2† | 17 (12 to 22)* | 15 (8 to 22)* |
| SpO<sub>2</sub>, % | 96.1 ± 0.4 | 93.0 ± 0.4 | -3.1 (-4.2 to -2.1)* | 97.2 ± 0.4 | 86.8 ± 0.4† | -10.4 (-11.5 to -9.4)* | -7.3 (-8.8 to -5.8)* |
| **Acute mountain sickness** |  |  |  |  |  |  |  |
| 2018 Lake Louise Score | 0.0 ± 0.3 | 0.3 ± 0.3 | 0.3 (-0.4 to 1.0) | 0.0 ± 0.3 | 1.9 ± 0.3† | 1.9 (1.2 to 2.6)* | 1.6 (0.6 to 2.6)* |
| 1993 Lake Louise Score | 0.0 ± 0.4 | 0.4 ± 0.4 | 0.4 (-0.6 to 1.4) | 0.0 ± 0.4 | 2.6 ± 0.4† | 2.6 (1.6 to 3.6)* | 2.2 (0.8 to 3.7)* |
| AMSc score | 0.00 ± 0.06 | 0.04 ± 0.06 | 0.04 (-0.12 to 0.21) | 0.00 ± 0.06 | 0.37 ±<br>0.06† | 0.36 (0.20 to 0.53)* | 0.32 (0.09 to<br>0.56)* |
| **Subjective sleep quality** |  |  |  |  |  |  |  |
| Karolinska Sleepiness Scale | 3.4 ± 0.3 | 4.0 ± 0.3 | 0.6 (-0.2 to 1.4) | 3.5 ± 0.3 | 3.8 ± 0.3 | 0.3 (-0.5 to 1.1) | -0.3 (-1.4 to 0.8) |
| Time until falling asleep, min | 17 ± 10 | 30 ± 10 | 13 (-13 to 38) | 14 ± 10 | 55 ± 10 | 41 (15 to 67)* | 27 (-9 to 65) |
| Number of nocturnal awakenings, n | 0.3 ± 0.3 | 0.8 ± 0.3 | 0.5 (-0.3 to 1.2) | 0.8 ± 0.3 | 1.7 ± 0.3 | 0.9 (0.2 to 1.6)* | 0.4 (-0.6 to 1.5) |
| Waketime after sleep onset, min | 2 ± 7 | 3 ± 7 | 2 (-18 to 21) | 3 ± 7 | 23 ± 7 | 20 (0 to 39) | 18 (-9 to 45) |
| Subjective sleep quality, % | 79 ± 4 | 75 ± 4 | -4 (-13 to 5) | 82 ± 4 | 58 ± 4† | -24 (-34 to -16)* | -20 (-33 to -8)* |

Values presented as means ± SE or mean differences (95%CI). \*P<0.05 versus 760 m. †p<0.05 vs. 3100 m. SpO<sub>2</sub>, arterial oxygenation assessed by finger pulse oximetry; AMSc, Environmental Symptom Questionnaire cerebral score.

## Discussion

This pilot study confirms that daily urinary sex hormone concentration monitoring using the commercially available fertility apps *Proov* and Mira is feasible for several consecutive days at high altitudes of up to 3,600 m, with the Proov device associated with lower error rates compared to *Mira*. Nevertheless, both hormone testing kits provide physiologically plausible urinary sex hormone concentrations to determine menstrual cycle phases and cycle days before, during, and after a high-altitude sojourn and can be considered as tools for future studies in premenopausal women evaluating susceptibility to AMS and its dependence on menstrual cycle phases. An important finding was that during the 2-day sojourns at high altitudes, the incidence of AMS was 43% at 3,100 m and 57% at 3,600 m. Moreover, other ARAHEs in the absence of AMS were observed at 3,600 m, encompassing fainting, fever, panic attacks, abdominal pain and diarrhoea. This underscores the diverse array of hypoxia-related, clinically relevant conditions in women that have rarely been reported.

### AMS and ARAHE

The comparably high incidence of AMS observed in our study is confirmed by generally accepted data. In a study by Li et al., during a two-day stay at an altitude of 3,270 m, women had a higher incidence of AMS than men (50% vs. 10%, p<0.05). Furthermore, multivariable regression analysis showed that not only the incidence, but also the severity of AMS, was significantly higher in women than in men (Li et al., 2022). In contrast, the 2023 UIAA statement on AMS in women suggests that there is no sex difference in AMS susceptibility; however, this statement has not been quantitatively confirmed by any meta-analysis examining the influence of sex on AMS. This situation, together with the revised LLS definition of AMS in 2018 and the lack of any recent prospective studies on AMS in women and men, does not allow evidence-based conclusions on sex-differences in AMS susceptibility.

Underlying sex-differences potentially contributing to the higher AMS susceptibility in women could be related to the lower hypoxic ventilatory response compared to men. In a study by Camacho-Cardenosa et al. (Camacho-Cardenosa et al., 2022), it was shown that, during the first hours of hypoxia, SpO_2_ decreased more markedly in women than in men, probably due to an initially lower and/or less efficient respiratory response to moderate hypoxia. In addition, there is evidence that women experience headaches more frequently at low altitudes (Li et al., 2023), indicating a potential susceptibility to AMS-like symptoms even before climbing to high altitude. This pre-existing condition may contribute to the development of AMS (MacGregor et al., 2006).

Adding even further variability, studies have reported differences in ventilation rates throughout the menstrual cycle (Loeppky et al., 2001; Rael et al., 2021; da Silva et al., 2006). In particular, PdG, acting on both peripheral and central chemoreceptors, is thought to be a potent respiratory stimulant (Bairam et al., 2015) enhancing minute ventilation during the mid-luteal phase compared to the follicular menstrual cycle phases (Bayliss and Millhorn, 1992; Beidleman et al., 1999). Our exploratory analysis showed the same correlation, indicating that higher PdG concentrations are associated with higher daytime SpO_2_ values (**Supplement Table S2**), independent of location.

Nevertheless, several previous studies and recommendations concluded that there are no sex-related differences in AMS susceptibility (Derstine et al., 2023b). The controversies will remain until we obtain robust findings from prospective studies specifically designed and powered to investigate and detect sex-related differences in AMS (defined by the revised LLS). With the emergence of wearables and modern technologies, this challenge become solvable, as demonstrated in our study that monitored hormones and determining menstrual cycle phases and cycle days of women staying at high altitudes. Our findings in premenopausal women are promising and important since they highlight the high adherence of participants monitoring urine hormones. *Proov*

### Limitations

This study was designed as a pilot study; therefore, due to the small sample size, the differences in the risk of AMS between menstrual cycle phases and altitudes that we found require corroboration in further studies. We applied a 30-day hormone-monitoring regimen; however fewer consecutive days of urine hormone monitoring might be sufficient to determine the menstrual cycle phases in participants staying at altitude. Despite the long duration of hormone monitoring, the observed error rate remained minimal and adherence of the participants high, suggesting that this approach is feasible in future studies. Absence of internet access may hamper immediate analysis of urinary hormone levels with the *Proov* kit. However, by being aware of such conditions, taking a photograph of the test strip noting the date and time allows for offline analysis at a convenient later time, which significantly reduces the risk of data loss.

## Conclusion

This pilot study in healthy, premenopausal women demonstrated good adherence and feasibility in monitoring female sexual hormones using the commercially available fertility monitoring kits and apps *Proov* and Mira during a sojourn at high altitudes. Premenopausal women showed a high AMS incidence at both 3,100 m and 3,600 m. The findings of this pilot study allow for evidence-based sample size estimations for future studies in women investigating AMS incidence and the efficacy of preventive AMS measures while monitoring and controlling for potential confounders such as sex hormones and menstrual cycle phases. For example, in a randomized, placebo-controlled parallel trial at 3,600 m investigating whether acetazolamide can reduce the incidence of AMS from 57% by 50% to 28.5%, a total of 94 women (48 per group), taking either acetazolamide or placebo, would be required for the final analysis, with 80% power and a two-sided alpha level of 5%.

## Data Availability

All data produced in the present study are available upon reasonable request to the authors.

## Acknowledgement

We gratefully acknowledge all participants for taking part in this study.

## Funding

This study was partly funded by the Swiss National Sciences Foundation (210361).

### Authorship confirmation statement

AT, MM, MF: Made substantial contributions to the conception and design of the work, investigation, acquisition, analysis, and interpretation of data for the work, writing – original draft, giving final approval of the version to be published, and agreeing to be accountable for all aspects of the work, ensuring that questions related to its accuracy or integrity are appropriately investigated and resolved. SU, AB, GM, MM, ML, CB, TMS, SU, KEB: Contributed significantly to the acquisition, analysis, and interpretation of data for the work, critically reviewed it for important intellectual content, gave final approval of the version to be published, and agreed to be accountable for all aspects of the work, ensuring that questions related to its accuracy or integrity are appropriately investigated and resolved.

## Abbreviations

AMS: Acute Mountain sickness
E1G: Etrone-1-glucuronide
PdG: Pregnanediol-3-alpha-glucuronide
LH: Luteinizing hormone
ARAHE: Altitude-related adverse health effects
LLS: Lake Louise score
SpO_2_: Oxygen saturation
PaCO_2_: Partial pressure of carbon dioxide
BMI: Body mass index
LLQ: Lake Louise questionnaires
UIAA: International Climbing and Mountaineering Federation

