## Supplemental Material for "Monitoring sexual hormones in women going to high altitude – a pilot study"

[*] *Shared first authorship*

***Funding***

This study was partly funded by the Swiss National Sciences Foundation (210361).

***Correspondence***

Taalaibekova Aijan

National Center of Cardiology and Internal Medicine,

Bishkek, Kyrgyzstan

**Supplement Table S1: Nocturnal pulse oximetry**

| Variable | 760 m | 1st night at 3,600 m | 2nd night at 3,600 m |
|---|---|---|---|
| Nocturnal $SpO_2$, % | 95.5 ± 0.6 | 79.3 ± 0.6* | 82.4 ± 0.6*# |
| Heart rate, bpm | 65 ± 2 | 85 ± 2* | 79 ± 2*# |
| Oxygen desaturation index, events/hour | 0.9 ± 0.5 | 8.6 ± 2.6* | 6.5 ± 0.6*# |
| $SpO_2$ <95%, % TIB | 15 ± 2 | 100 ± 2* | 100 ± 2* |
| $SpO_2$ <90%, % TIB | 1 ± 1 | 99 ± 1* | 98 ± 1* |
| $SpO_2$ <85%, % TIB | 0 ± 4 | 94 ± 4* | 69 ± 4*# |
| $SpO_2$ <80%, % TIB | 0 ± 5 | 48 ± 6* | 20 ± 6*# |
| $SpO_2$ <75%, % TIB | 0 ± 4 | 13 ± 4* | 5 ± 4 |
| $SpO_2$ <70%, % TIB | 0 ± 1 | 3 ± 1 | 0 ± 1 |

Values presented as means ± SE. *p<0.05 compared to 760 m; #p<0.05 2nd night versus 1st night at 3,600 m. $SpO_2$, arterial oxygenation assessed by finger pulse oximetry; TIB, time in bed.

**Supplement Table S2. The effect of sex hormones and altitude on arterial oxygenation – a mixed linear regression model**

| Dependent variable: Daytime arterial oxygenation by finger pulse oximetry, % | **Coefficient** | **Standardized beta coefficient** | **Std. Error** | **P-value** | **95% CI** |
|---|---|---|---|---|---|
| **Progesterone concentration, mcg/ml** | 0.17 | 0.21 | 0.04 | <0.001 | 0.09 to 0.25 |
| **Estrogen, ng/ml** | -0.01 | -0.13 | 0.00 | 0.007 | -0.01 to 0.00 |
| **Luteinizing hormone, mlU/ml** | 0.00 | 0.00 | 0.02 | 0.962 | -0.03 to 0.03 |
| **Altitude of assessment** | | | | | |
| 760 m (*reference*) | 96.1 | NA | 0.4 | <0.001 | 95.3 to 96.8 |
| Difference between 3,100 and 760 m | -3.3 | -0.32 | 0.5 | <0.001 | -4.3 to -2.2 |
| Difference between 3,600 and 760 m | -9.8 | -0.95 | 0.6 | <0.001 | -10.9 to -8.6 |

Mixed linear regression analysis including all available measurements. Due to the small sample size, the parameters for the mixed models were predefined. For example, according to this model, a 1 mcg/ml increase in progesterone is associated with a 0.17% increase in $SpO_2$, independent of altitude and other hormones; an altitude sojourn at 3,600 m is associated with a 9.8% decrease in $SpO_2$, independent of the hormone levels.
